# Trends in the Distribution of *P Values* in Epidemiology Journals: Decreased *P* Hacking or Increased Power?

**DOI:** 10.1101/2025.02.13.25322235

**Authors:** Sarah F. Ackley, Ryan M. Andrews, Christopher Seaman, Michael Flanders, Ruijia Chen, Jingxuan Wang, Grissel Lopes, Kendra D. Sims, Peter Buto, Erin Ferguson, Isabel Elaine Allen, M. Maria Glymour

## Abstract

Epidemiologists have advocated for reporting confidence intervals and deemphasizing *P* values to address long-standing concerns about null-hypothesis statistical-significance testing, *P* hacking, and reproducibility. It is unknown if efforts to reduce reliance on *P* values have altered the distribution of *P* values. For 21,377 abstracts published 2000-2024 in 4 major epidemiology journals, two-sided *P* values (N=25,360) were calculated from estimates and confidence intervals scraped using ChatGPT’s 4o model. We evaluated trends over time to determine whether the empirical distribution of *P* values changed. We fitted to expected *P* value distributions and simulated these distributions with and without assuming changes in statistical power over time. Average *P* values decreased from 2000 to 2024, as did the fraction just below the 0.05 threshold. Fits to models indicate that statistical power increased. Increasing power would reduce average *P* value while also decreasing *P* values near the 0.05 threshold—precisely the trends observed in epidemiology journals. Although the frequency of *P* values near the 0.05 threshold has declined modestly, this likely reflects increases in statistical power rather than decreases in *P* hacking.

## Introduction

There have been long-standing concerns about null-hypothesis statistical-significance testing,^1–4^ and, over the last few decades, substantial critiques have emerged regarding statistical education and the use of *P* values in the scientific literature.^5,6^ *P* values are widespread in epidemiological and clinical research,^7^ yet it has been argued that their overemphasis contributes to *P* hacking and the underreporting of non-statistically significant results.^8^ In this context, the positive predictive value of a reported statistically significant association for a true association in the target population may be low, contributing to the reproducibility crisis.^9,10^ Epidemiologists and epidemiology journals have played a prominent role in arguing for an increased emphasis on reporting confidence intervals to foster rigorous statistical thinking and reduce misinterpretation of findings.^1,11,12^ Some epidemiologists have gone further to argue that *P* values be omitted except in select circumstances.^7,13^ Epidemiology journals discourage overreliance on *P* values and expressly favor confidence intervals over *P* values (Table S1).^13^

If changing scientific norms and policies have limited discussion and reporting of *P* values,^1,13–16^ this may have curtailed *P* hacking and ameliorated publication bias based on statistical significance.^5^ We might only expect to see changes in the distribution of *P* values recently, as those who were trained during the reproducibility crisis^10,17,18^ entered the research workforce.

However, it is also plausible that the impact is negligible since the problems associated with *P* values are inherent to frequentist analysis: confidence intervals (CIs), which are typically constructed to be congruent with *P* values,^19^ can easily be used to engage in *P* hacking by determining analyses that result in an CI excludes the null.^1,6^

Other trends may impact *P* hacking and publication bias. In the context of multiple decision points in an analysis, *P* hacking need not be intentional to occur.^20,21^ It is plausible that unintentional or intentional *P* hacking is more likely to occur under growing pressures to publish.^22^ As collecting data becomes cheaper and easier,^17^ the growing availability of datasets with large numbers of variables may make it easier to *P* hack since there are more possible decision points.^23^ Finally, since epidemiologists and epidemiology journals were early adopters of the CI-only reporting approach,^24^ changes in *P* hacking may have been minimal since the year 2000.

We evaluate trends in *P* values derived from confidence intervals since 2000 in abstracts at *Epidemiology*, the *American Journal of Epidemiology* (AJE), the *European Journal of Epidemiology* (EJE), and the *International Journal of Epidemiology* (IJE). We compare naive regression results to results from *P*-curve analyses that build on methods in the social psychology literature. These methods were developed to evaluate growing concerns about research fraud and reproducibility in the field. Using *P-*curve methods, we evaluate the plausibility that the movement to reduce reliance on *P* values has resulted in changes in the distribution of derived *P* values since 2000.

## Methods

A schematic of the analysis is shown in figure 1.

**Figure 1:**
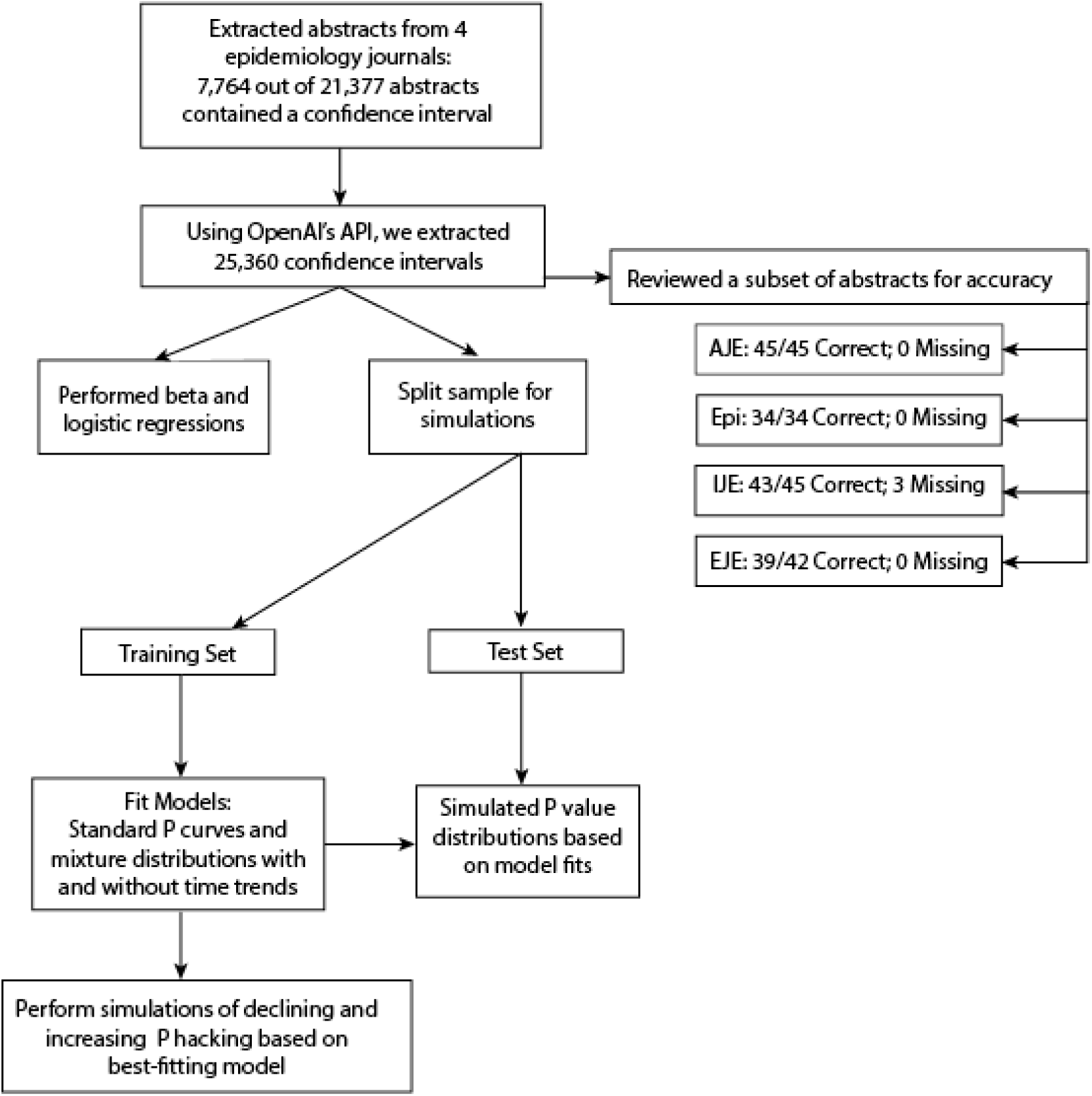
Flowchart of the Analysis.

### Data Extraction

A comprehensive list of abstracts from 1/1/2000 to 12/31/2024 from *Epidemiology*, AJE, EJE, and IJE was obtained from PubMed (pubmed.ncbi.nlm.nih.gov). Search strings are given in the supplemental methods. Abstracts were downloaded as a text file and split into individual files using Python3 (version 3.9.6). Based on example extractions and narrative instructions, OpenAI’s API with the ChatGPT 4o engine was used to extract estimates, lower confidence bounds, and upper confidence bounds for each abstract, along with date of publication, journal, and pubmed ID. Each estimate and confidence interval pair was written to file with the PubMed ID, date of publication, and whether the estimate was likely a ratio measure or linear regression coefficient.

R (version 4.3.2) was used for all statistical analysis, model fitting, and simulation. Approximate two-sided *P* values were calculated using the following:

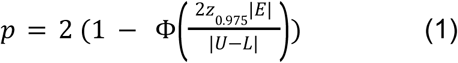

where 𝑝 is the calculated approximate *P* value, Φ is the cumulative density function of the normal distribution, |𝐸| is the absolute values of estimate, |𝑈 − 𝐿| is the length of the confidence interval (the absolute value of the upper limit minus the lower limit), and 𝑧_0.975_ ≈ 1. 96 is the standard normal deviate. Measures on the ratio scale, as determined by the text preceding the estimate in the abstract, were logarithmized prior to calculating the *P* value.

### Naive Regression Analyses

Beta regression with a logit link was used to evaluate changes in *P* values over time and logistic regression was used to evaluate changes in the odds of a *P* value being on the intervals (0.01, 0.05], (0.03,0.05], (0,0.05], and (0,0.01]. Mixed-effects beta and logistic regressions with random intercepts by article were also estimated. We estimated models for overall time trends adjusted for journal and models stratified by journal.

### *P-C*urve Analyses

We contextualized and interpreted changes in *P* values over time drawing on existing literature on *P* curves.^25–28^ *P* value distributions (curves) under the null hypothesis follow a uniform distribution; *P* curves under an alternative hypothesis follow a monotonic curve with density increasing without bound as *P* approaches zero.^25^ In simple settings where all hypotheses are equally powered, this curve is parameterized by a single quantity, which we will denote θ; this is sometimes referred to as a Cohen’s *d* in the literature, but we do not adopt this term because the term Cohen’s *d* is used for other quantities. This parameter θ is the ratio of the absolute value of the true effect over its standard error. Statistical power is a function of θ and is given by the following: 𝑃 = 1 + Φ(− 𝑧_0.975_ − θ) − Φ(𝑧_0.975_ − θ), where 𝑃 is the statistical power and assuming two-sided *z*-statistics. Thus, θ can be thought of as a transform of the statistical power.

The cumulative distribution function of two-sided *P* values as a function, where two-sided *P* values are calculated for equally powered hypotheses and assuming *z*-statistics is given by the following:

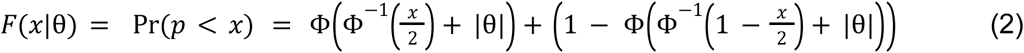

where 𝐹(𝑥) is the probability that a *P* value is less than some quantity 𝑥 ∈ (0, 1) and 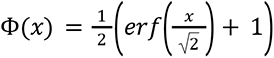 is the cumulative density function of the normal distribution.

Taking the derivative of the cumulative distribution function (𝑥) in equation (2), we have that the probability density function 𝑓(𝑥) is given by the following:

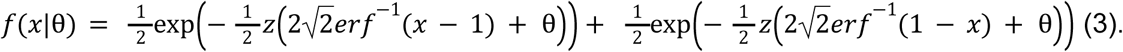

Symbolic differentiation and simplification was checked using numeric differentiation in R. Plugging in to equation 2, when θ = 0, 𝐹(𝑥) = 𝑥/2 + 𝑥/2 = 𝑥, which implies that the probability density function 𝑓(𝑥) is uniform over 𝑥 ∈ (0, 1) if the null hypothesis is true, as is well known.

We used maximum-likelihood estimation to fit the probability density function in equation (3) to the empirical *P* value data extracted from epidemiology journals to identify the best fitting estimate of the standardized effect size θ of underlying studies. Equation 3 describes the expected distribution under the narrow assumption that all studies presented in the journals had the same θ and thus statistical power. We also fit the data to expected distributions under more flexible assumptions. First, we assumed all studies were either null or had θ of θ_2_(where θ_2_ is estimated from the data). This mixture distribution is generated with two unequally weighted Dirac δ-functions. We also fit the observed *P* values to the distribution assuming θ follows an exponential distribution and a gamma distribution.

We next used maximum likelihood to evaluate time trends in the *P* value distribution. To do this, we incorporated a slope *m* into the *P*-value distribution equation and fit the maximum likelihood model to the observed P value data, defining the likelihood as: 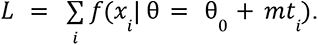 For the time-varying version of the two Dirac δ-function mixture model, we have θ_2_increasing with time while the fraction of null hypotheses remains constant. For the time-varying version of the exponential-mixture model, the mean increases linearly with time. For the time-varying version of the gamma-mixture model, the mean increases linearly with time by allowing the shape parameter to increase linearly with time while keeping the scale parameter constant. We compare model fits using the Akaike information criterion.

We evaluate the plausibility of time trends in the distribution of *P* values based on fitted models. Prior to fitting, we split the data into equally sized training and testing sets. Fitting was performed using the training set data for each of the above distributions. Expected *P* value distributions were then simulated based on the fitted coefficients and the dates of publication represented in the testing data set. The expected distributions were then each compared to the observed distributions in the testing set data. Expected fractions were also calculated based on the dates of publication in the testing data using parameters fit in the training set. Specifically, cumulative distribution functions for each of the fits were used to estimate expected fractions for the following intervals considered potentially indicative of *P* hacking: (0,0.05]; (0,0.01]; (0.01,0.05]; and (0.03,0.05]. Simulated values from each probability density function were obtained by generating uniform random variables and using the probability integral transform.

Specifically, for each random value drawn from the standard uniform distribution, the cumulative distribution function for the *P* value distribution is set equal to that value and solved for the corresponding *P* value. One hundred simulations were performed for each model.

### Simulations of *P* Hacking

The determine expected *P* value distributions generated by constant *P* hacking or time trends in *P* hacking, we simulated P value distributions under three scenarios from the best model fit without time dependence of θ: First, we simulated a scenario where *P* hacking and statistical power remain constant over time. We conceptualize *P* hacking as occurring if the researcher conducts repeated analyses until finding a *P* value below some threshold or reaches a maximum number of attempts. We generated a *P* value distribution under *P* hacking by iterating the following steps a number of times equal to the total number of *P* values:

1. Select a maximum number of attempts drawing from a Poisson distribution with a mean of 5.
2. Generate a θ using the parameters from the best model fit (model 4).
3. Generate a *P* value using the cumulative distribution function of the *P* value distribution for a single effect size and the probability integral transform.
4. If the generated *P* value was less than 0.05, accept it; otherwise, generate a new *P* value until the maximum number of attempts is reached. If no P value is below 0.05, only the return only the final *P* value.

Next, we simulated a scenario where *P* hacking decreases with time and statistical power remains constant over time. In this scenario, the mean of the Poisson distribution used to generate attempts decreases over time (5 at the midpoint of 2012 and changing by -0.4 per year). The same four steps were applied, and publication times from the original sample were used to generate mean θ as a function of time. Next, we simulated a scenario where *P* hacking increases with time and statistical power remains constant over time. In this scenario, the mean of the Poisson distribution used to generate attempts decreases over time (5 at the midpoint of 2012 and changing by 0.4 per year). 100 simulations of sets of *P* values were generated for each scenario; beta and logistic regression models were fit to the simulated data.

## Results

### Data Extraction

As shown in Figure and Table 1, 25,360 *P* values were calculated from 7,764 abstracts that contained at least one confidence interval, out of 21,377 abstracts in total. Automated extraction performed well with respect to manual extraction from random subsamples of 25 abstracts per journal: error rates were Epidemiology 0%, 95% CI: (0%, 10.2%); AJE 0%, 95% CI: (0%, 7.8%); EJE 7.1%, 95% CI: (1.5%, 19.4%), IJE 4.4%, 95% CI: (0.5%, 15.1%). Out of over 25,000 confidence intervals, 78 entries were dropped at the logarithmizing step due to entries less than or equal to zero in the estimate or confidence limits. A histogram of P values is given in Figures 1A (all data) and 1B (training data) along with the median in each journal in each year in Figure 1C.

**Table 1:** Number of abstracts and confidence intervals and summary statistics by journal and overall. Table S2 gives summary statistics by both journal and year. Epi=*Epidemiology*, AJE=*American Journal of Epidemiology*, EJE=*European Journal of Epidemiology*, IJE=*International Journal of Epidemiology.* IQR: interquartile range; *N*: number; CI: confidence interval.

| Journal | *N* of Abstracts | *N* of CIs | Mean of *P* | Median of log *P* (IQR) |
| --- | --- | --- | --- | --- |
| Epi | 1454 | 4530 | 0.102 | -4.85, (-12.5, -2.88) |
| AJE | 3475 | 10577 | 0.0891 | -5.11, (-12.4, -3.14) |
| EJE | 1092 | 3882 | 0.0742 | -5.9, (-16.6, -3.42) |
| IJE | 1743 | 6371 | 0.0734 | -6.49, (-20.5, -3.45) |
| Overall | 7764 | 25360 | 0.0853 | -5.45, (-14.8, -3.19) |

### Naive Regression

The fixed-effects beta regression models in the full data suggest that overall *P* values decreased over time (Table 2; Table S3 results stratified by journal). Specifically, the odds of *P* decreased by 16.9% per decade (ratio = 0.831, 95% CI: (0.814, 0.847)). The logistic regression models indicated that the odds of a *P* value falling at or below 0.05 increased by 24% per decade (OR=1.24, 95% CI: (1.19, 1.29)) and the odds of a *P* value falling at or below 0.01 increased by 31% per decade (OR=1.31, 95% CI: (1.27, 1.36)). The odds of a *P* value falling *between* 0.01 and 0.05 decreased by 16.3% per decade (OR=0.837, 95% CI: (0.801, 0.874)) and the odds of a *P* value falling between 0.03 and 0.05 decreased by 14.1% per decade (OR=0.859, 95% CI: (0.803, 0.92)). Mixed-effects regression models indicated similar time trends.

**Table 2:**
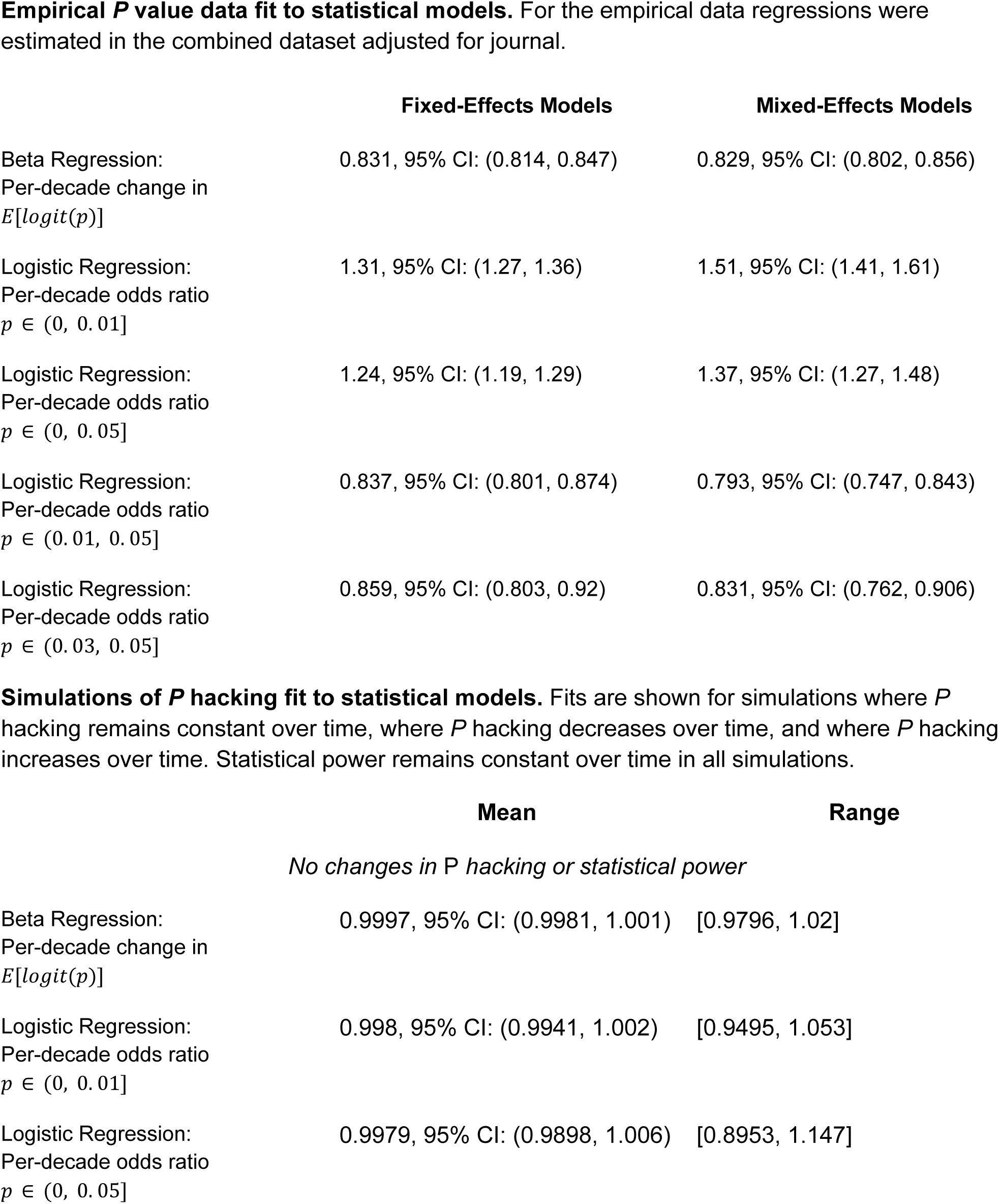
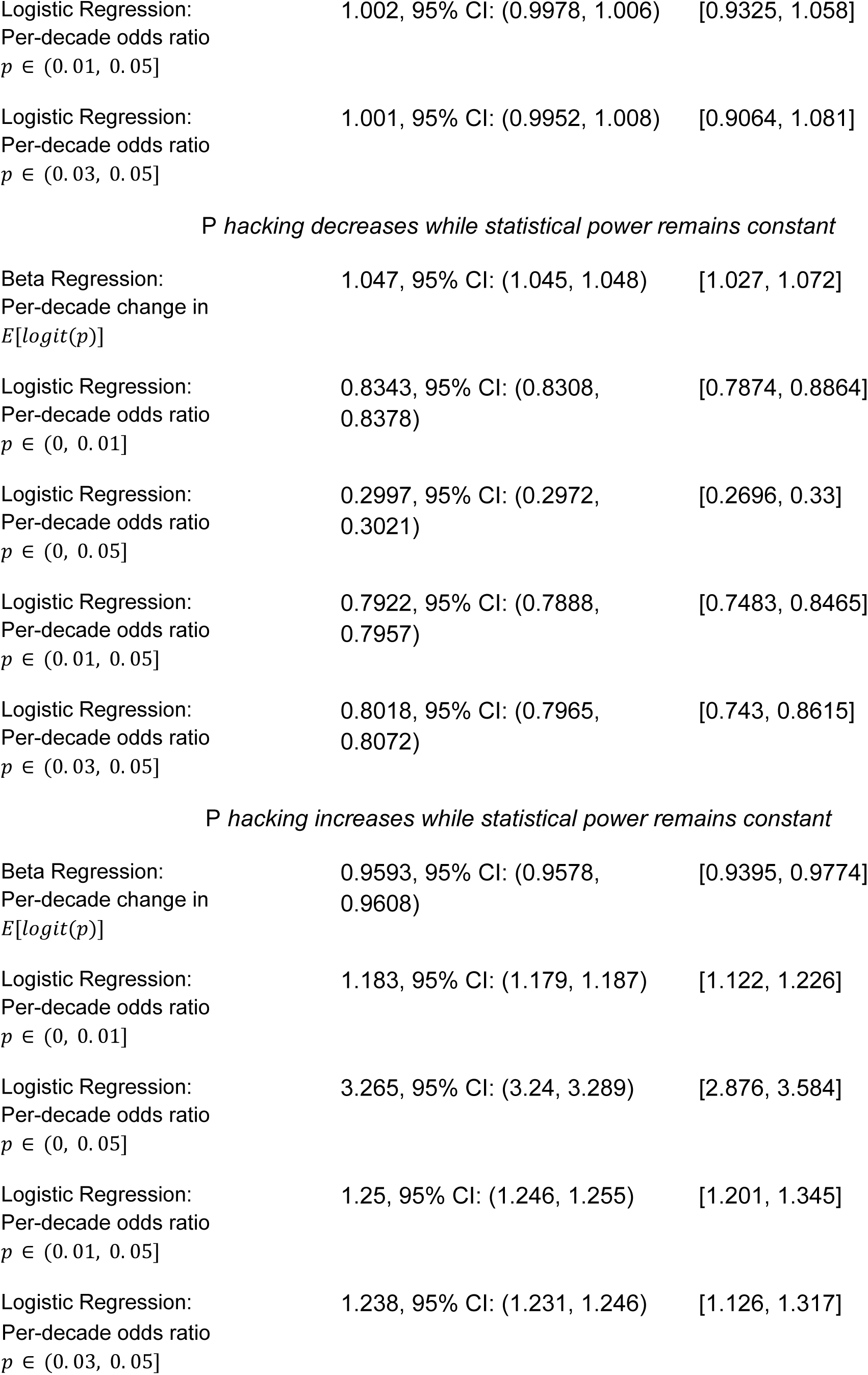
Regression coefficients from beta- and logistic-regression models estimating per-decade change in features of empirical and simulated P value distributions.

### *P*-Curve Analyses

Table 3 gives estimated model parameters fitted to the observed data under 8 possible distribution families for *P* values and Figure 1B shows the model fits overlaid on the testing data of observed *P* values. Models where θ increases over time and mixture models perform better than models with time-constant θ or a single θ; the gamma mixture model with a linear time-trend outperformed other models. Log likelihoods and Akaike information criteria are given in the Supplement.

**Table 3:** Estimated parameters under 8 possible rules for generating *P*-values. All estimates are rounded to 3 significant figures without trailing zeros.

| Model | Parameters | Estimate (95% CI) | Percent Power (95% CI) |
| --- | --- | --- | --- |
| 1. All *P* values generated with a single $\theta$ (effect size divided by standard error) | $\theta$ | 3.37, 95% CI: (3.35, 3.39) | 92.1, 95% CI: (91.8, 92.3) |
| 2. A single $\theta$ with a linear time trend | $\theta$ at 1/1/2012 | 3.35, 95% CI: (3.33, 3.37) | 91.8, 95% CI: (91.5, 92.1) |
|  | Time trend | .0355, 95% CI: (.033, .0379) |  |
| 3. Two θs ($\theta_1=0$ and $\theta_2$) | Mean $\theta$ | 3.06, 95% CI: (3.02, 3.09) | 86.3, 95% CI: (85.5, 87.1) |
|  | Proportion null | .255, 95% CI: (.245, .265) |  |
| 4. Two θs ($\theta_1=0$ and $\theta_2$) with a linear time trend | Mean $\theta$ at 1/1/2012 | 3.04, 95% CI: (3, 3.08) | 86, 95% CI: (85.2, 86.8) |
| | Proportion $\theta_1=0$ | .253, 95% CI: (.243, .263) | |
|  | Time trend | .0366, 95% CI: (.034, .0392) |  |
| 5. Exponential mixture ($\theta$ is sampled from an exponential distribution) | Mean $\theta$ | 3.14, 95% CI: (3.09, 3.2) | 88.2, 95% CI: (87, 89.3) |
| 6. Exponential mixture with a linear time trend | Mean $\theta$ at 1/1/2012 | 3.12, 95% CI: (3.06, 3.18) | 87.7, 95% CI: (86.5, 88.8) |
|  | Time trend | .0371, 95% CI: (.0289, .0453) |  |
| 7. Gamma mixture ($\theta$ is sampled from an gamma distribution) | Mean $\theta$ | 3.35, 95% CI: (3.32, 3.39) | 91.8, 95% CI: (91.3, 92.4) |
|  | Scale | .949, 95% CI: (.914, .985) |  |
| 8. Gamma mixture, linear trend | Mean $\theta$ at 1/1/2010 | 3.33, 95% CI: (3.3, 3.37) | 91.5, 95% CI: (90.9, 92.1) |
|  | Time trend | .0358, 95% CI: (.033, .0386) |  |
|  | Scale | .927, 95% CI: (.891, .962) |  |

The expected fraction of *P* values falling within specific ranges potentially indicative of *P* hacking based on simulations from fitted models are given in Table 4, with comparison to the fraction of *P* values in each range actually observed in the journals. For all models, there is an excess of *P* values between 0.01 and 0.05 and between 0.03 and 0.05 compared with fractions expected based on simulations. None of the 100 simulations produced fractions in these ranges as large as observed in major epidemiology journals. Simulations indicate that there are more *P* values greater than 0.05 than would be expected from models 1 and 2, but for models 3-8 there are fewer than would be expected.

**Table 4:**
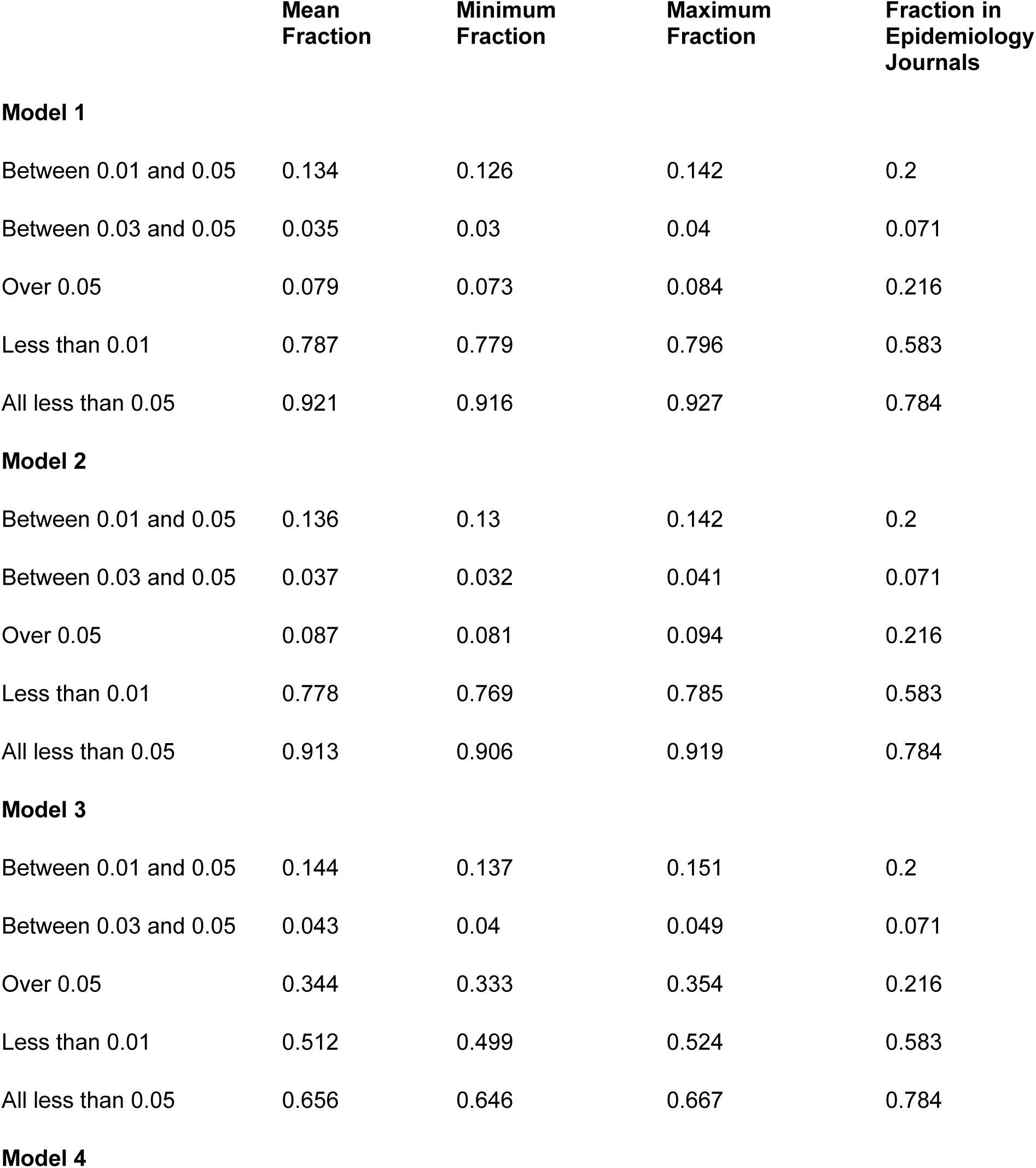
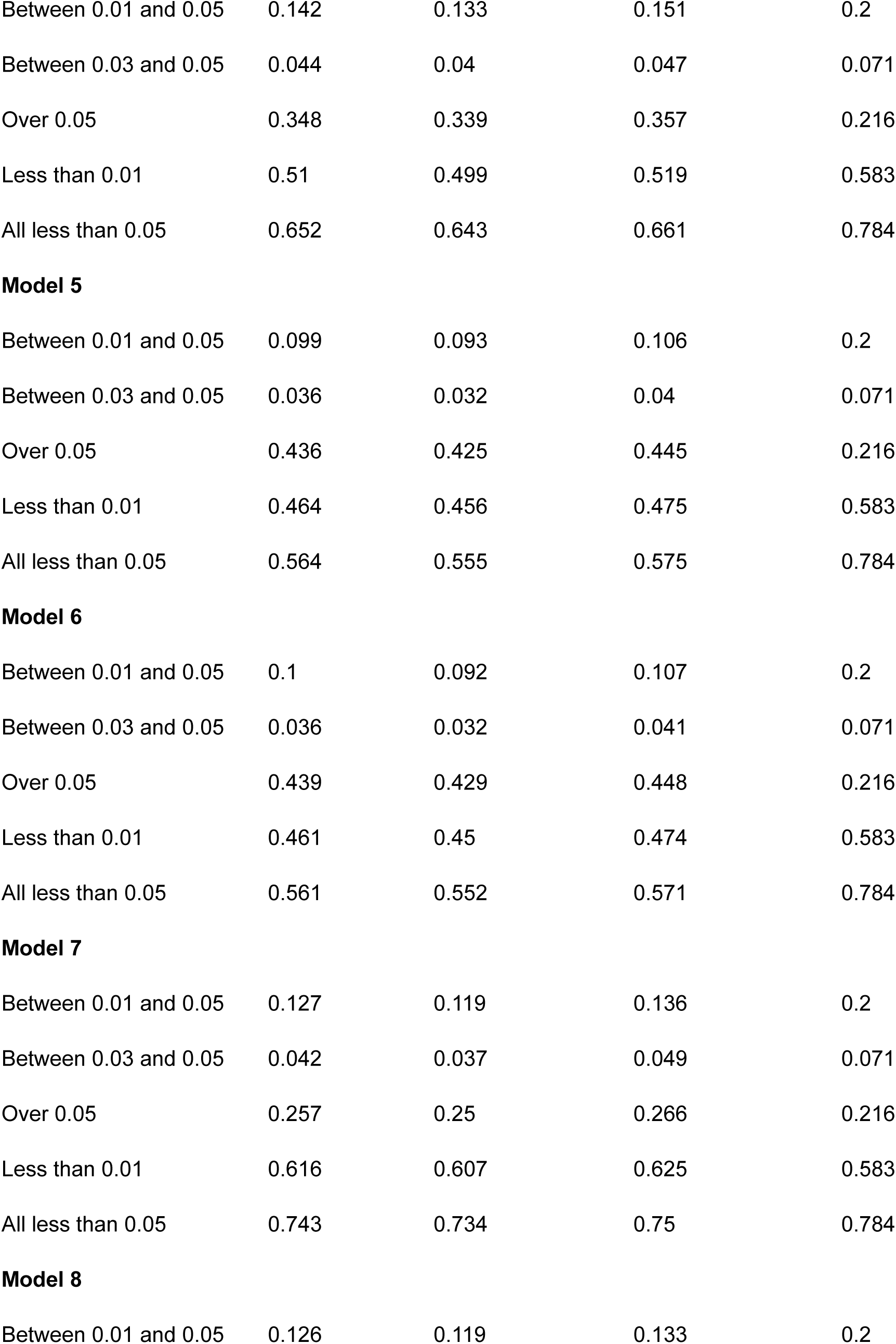
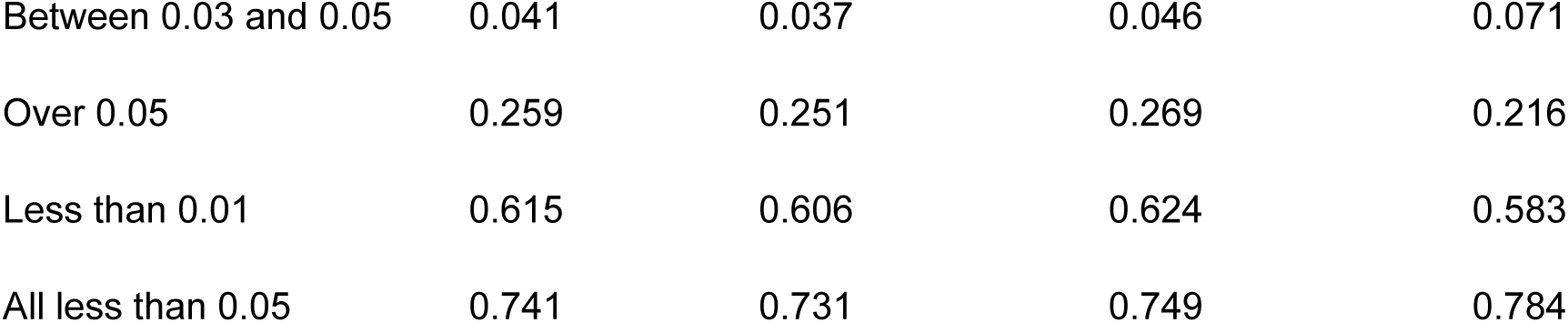
Simulated fractions compared with fractions in the epidemiology literature. Models are as follows: 1. One θ; 2. One θ, linear trend; 3. Two θs (θ_1_=0 and θ_2_); 4. Two standardized effect sizes (θ_1_=0 and θ_2_), linear trend; 5. Exponential mixture; 6. Exponential mixture, linear trend; 7. Gamma mixture; and 6. Gamma mixture, linear trend. Model parameters are from fits to the training set, while simulation abstract dates and the fractions in AJE are from the testing set.

|  | Mean Fraction | Minimum Fraction | Maximum Fraction | Fraction in Epidemiology Journals |
| --- | --- | --- | --- | --- |
| **Model 1** |  |  |  |  |
| Between 0.01 and 0.05 | 0.134 | 0.126 | 0.142 | 0.2 |
| Between 0.03 and 0.05 | 0.035 | 0.03 | 0.04 | 0.071 |
| Over 0.05 | 0.079 | 0.073 | 0.084 | 0.216 |
| Less than 0.01 | 0.787 | 0.779 | 0.796 | 0.583 |
| All less than 0.05 | 0.921 | 0.916 | 0.927 | 0.784 |
| **Model 2** |  |  |  |  |
| Between 0.01 and 0.05 | 0.136 | 0.13 | 0.142 | 0.2 |
| Between 0.03 and 0.05 | 0.037 | 0.032 | 0.041 | 0.071 |
| Over 0.05 | 0.087 | 0.081 | 0.094 | 0.216 |
| Less than 0.01 | 0.778 | 0.769 | 0.785 | 0.583 |
| All less than 0.05 | 0.913 | 0.906 | 0.919 | 0.784 |
| **Model 3** |  |  |  |  |
| Between 0.01 and 0.05 | 0.144 | 0.137 | 0.151 | 0.2 |
| Between 0.03 and 0.05 | 0.043 | 0.04 | 0.049 | 0.071 |
| Over 0.05 | 0.344 | 0.333 | 0.354 | 0.216 |
| Less than 0.01 | 0.512 | 0.499 | 0.524 | 0.583 |
| All less than 0.05 | 0.656 | 0.646 | 0.667 | 0.784 |
| **Model 4** |  |  |  |  |
| Between 0.01 and 0.05 | 0.142 | 0.133 | 0.151 | 0.2 |
| Between 0.03 and 0.05 | 0.044 | 0.04 | 0.047 | 0.071 |
| Over 0.05 | 0.348 | 0.339 | 0.357 | 0.216 |
| Less than 0.01 | 0.51 | 0.499 | 0.519 | 0.583 |
| All less than 0.05 | 0.652 | 0.643 | 0.661 | 0.784 |
| **Model 5** |  |  |  |  |
| Between 0.01 and 0.05 | 0.099 | 0.093 | 0.106 | 0.2 |
| Between 0.03 and 0.05 | 0.036 | 0.032 | 0.04 | 0.071 |
| Over 0.05 | 0.436 | 0.425 | 0.445 | 0.216 |
| Less than 0.01 | 0.464 | 0.456 | 0.475 | 0.583 |
| All less than 0.05 | 0.564 | 0.555 | 0.575 | 0.784 |
| **Model 6** |  |  |  |  |
| Between 0.01 and 0.05 | 0.1 | 0.092 | 0.107 | 0.2 |
| Between 0.03 and 0.05 | 0.036 | 0.032 | 0.041 | 0.071 |
| Over 0.05 | 0.439 | 0.429 | 0.448 | 0.216 |
| Less than 0.01 | 0.461 | 0.45 | 0.474 | 0.583 |
| All less than 0.05 | 0.561 | 0.552 | 0.571 | 0.784 |
| **Model 7** |  |  |  |  |
| Between 0.01 and 0.05 | 0.127 | 0.119 | 0.136 | 0.2 |
| Between 0.03 and 0.05 | 0.042 | 0.037 | 0.049 | 0.071 |
| Over 0.05 | 0.257 | 0.25 | 0.266 | 0.216 |
| Less than 0.01 | 0.616 | 0.607 | 0.625 | 0.583 |
| All less than 0.05 | 0.743 | 0.734 | 0.75 | 0.784 |
| **Model 8** |  |  |  |  |
| Between 0.01 and 0.05 | 0.126 | 0.119 | 0.133 | 0.2 |
| Between 0.03 and 0.05 | 0.041 | 0.037 | 0.046 | 0.071 |
| Over 0.05 | 0.259 | 0.251 | 0.269 | 0.216 |
| Less than 0.01 | 0.615 | 0.606 | 0.624 | 0.583 |
| All less than 0.05 | 0.741 | 0.731 | 0.749 | 0.784 |

Figure 2A extends results from Table 4 to show changes over time in the expected fractions of *P* values within intervals potentially indicative of *P* hacking derived from the model fits to the training data. Fractions for the testing dataset are plotted by year. The fraction of observed *P* values between 0.01 and 0.05 and between 0.03 and 0.05 exceed the expected fractions produced by all models for every year from 2000 to 2024. Specifically, the observed distribution indicates an excess of *P* values at or just below the 0.05 threshold compared to any of the expected distributions. However, trends in fractions of *P* values within each interval increase over time, similar to the expected trends from models 2 (two effect sizes), 4 (exponential mixture), and 6 (gamma mixture). From 2000 to 2024, the fraction of published P values below 0.01 increased while the fraction between 0.03 and 0.05 decreased, consistent with model expectations.

**Figure 2:**
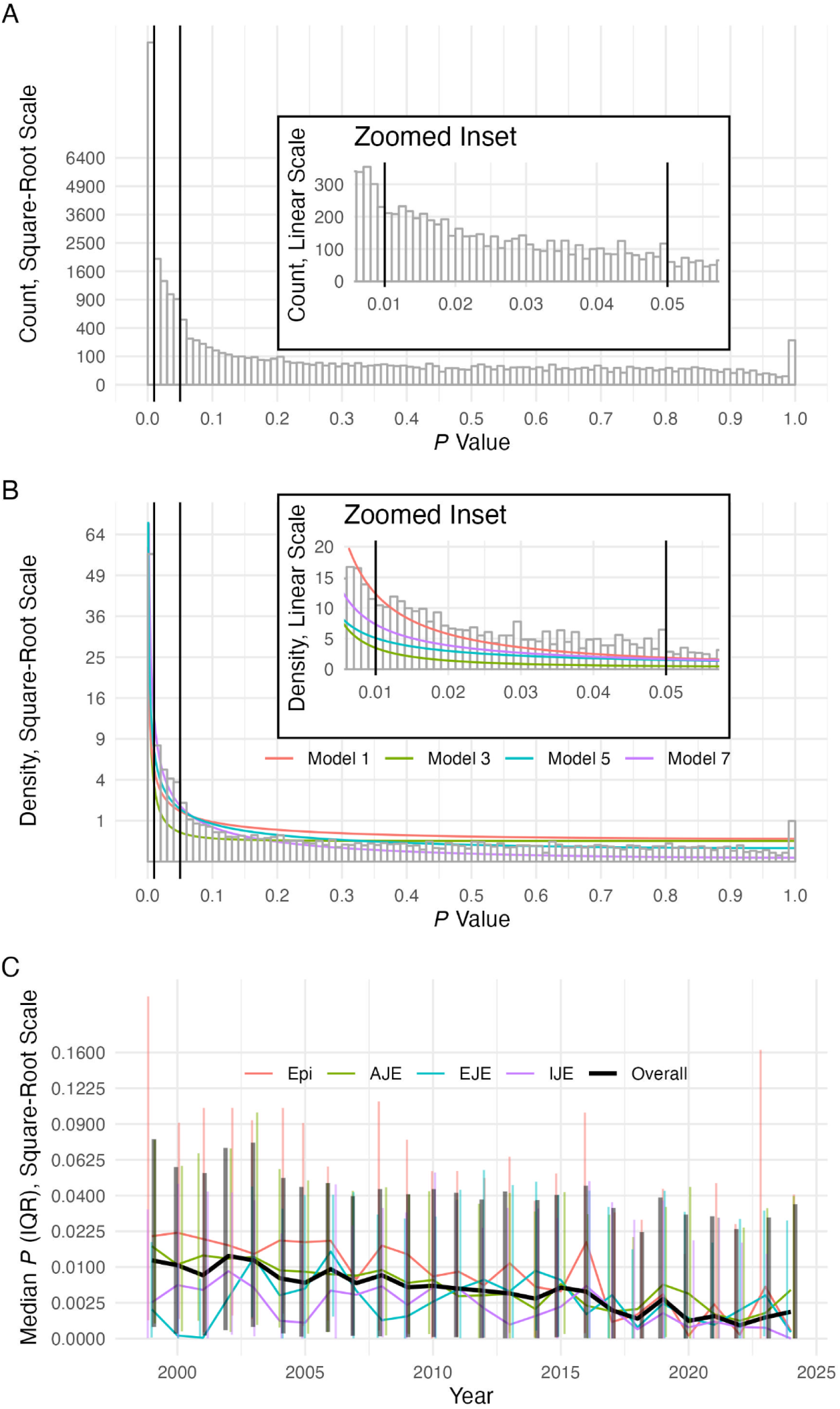
Distribution of *P* values at AJE since 2000 and model fits. **A.** Counts of P values in the full dataset. **B.** Density of P values for the training set and model fits for odd-numbered models. *P* value density is a function of time for even-numbered models and thus they are not plotted. For both **A** and **B,** vertical bars denote 0.01 and 0.05. **C.** Median and interquartile range (IQR) of *P* values over time, showing *P* values are declining over time.

**Figure 3:**
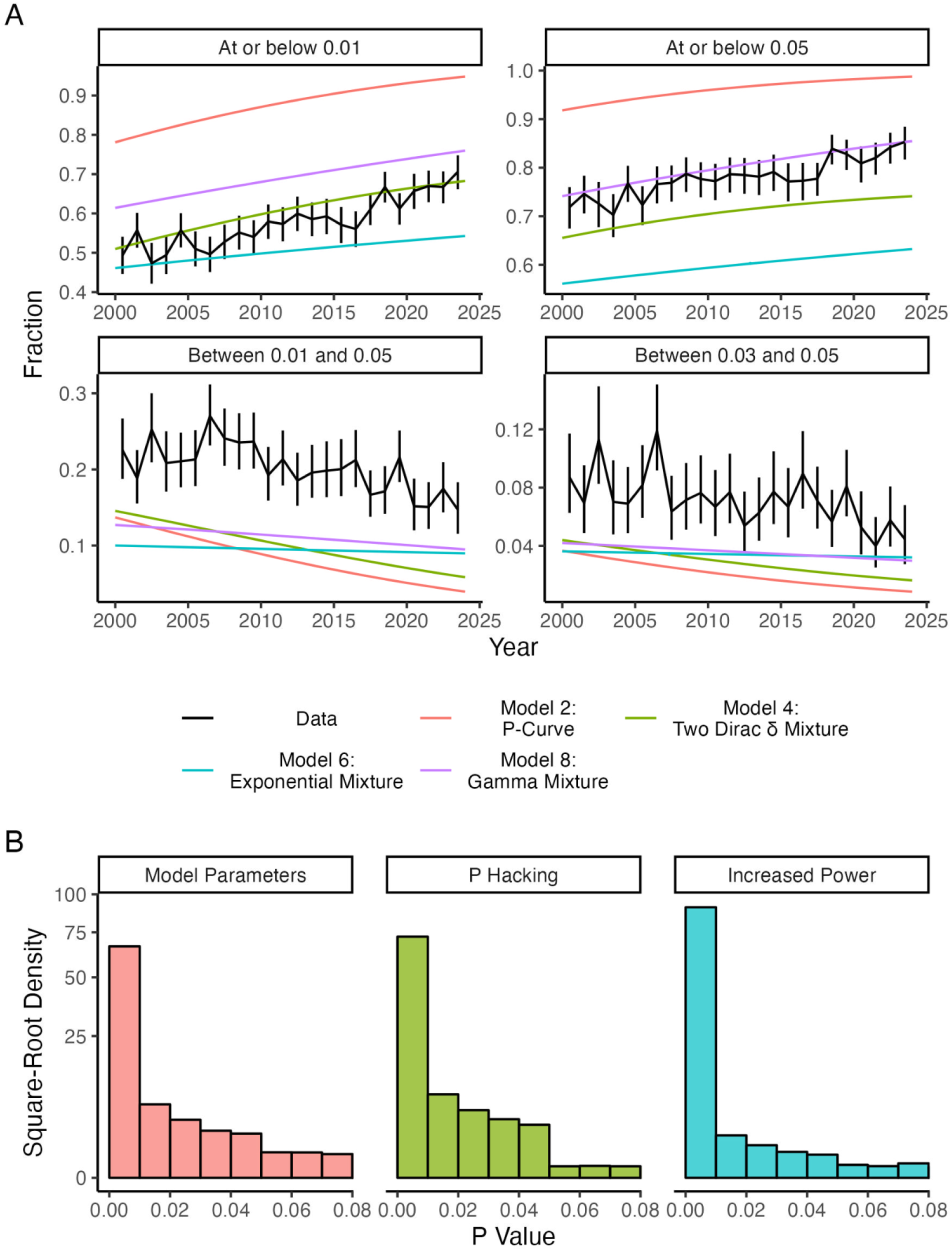
Expected trends in fractions of *P* values and simulated *P* Values. **A.** Expected fractions of *P* values falling within ranges potentially indicative of *P* hacking, predicted from 2000 through 2024 under 4 possible models with time-varying parameters, compared with fractions (exact binomial confidence intervals). Expected and summaries of simulated fractions of *P* values are plotted for the following time-varying models: 2. One standardized effect size (θ), linear trend; 4. Two standardized effect sizes (θ*_1_*=0 and θ*_2_*), linear trend; 6. Exponential mixture, linear trend; and 8. Gamma mixture, linear trend. All *P* values at or below 0.01 and 0.05 are expected to increase, while *P* values between 0.01 and 0.05 and between 0.03 and 0.05 are expected to decrease. As in Figure 1, none of the models fit the data well for *P* values ranging from 0.01 and 0.05 and 0.03 and 0.05. **B.** 25,360 simulated *P* values for three scenarios. Model Parameters: best-fitting gamma-mixture model without modification. *P* Hacking: the gamma-mixture model with the addition of *P* hacking, wherein *P* values over 0.05 are redrawn up to 5 times. Increased Power: the gamma-mixture model with statistical power increased by doubling the scale parameter. P hacking removes density from above 0.05 and moves it largely between 0.01 and 0.05, with only small increases in the density of *P* values below 0.01. Increasing statistical power increases density below 0.01, while removing density in the 0.01 to 0.05 range.

### Simulations of *P* Hacking

Table 2 and Figure 2B show the expected annual changes in the distribution of *P* values under assumed time-constant *P* hacking, decreasing *P* hacking, or increasing *P* hacking, all with time-constant statistical power. The simulation results in Table 5 indicate the empirically observed trends in *P* value distribution were unlikely to occur without changes in statistical power, i.e., they cannot be explained simply by changes in *P* hacking. Specifically, when there is change in neither *P* hacking nor statistical power, estimated coefficients for the time trends from beta- and logistic-regression models are on average very near unity—indicating no temporal changes—and inconsistent with the observed time trends shown in Table 2 columns 1 or 2.

**Table 5:** Table synthesizing qualitative trends based on statistical models and simulations. The empirically observed trends match the scenario where statistical power is increasing; the corresponding rows have been highlighted.

| Scenario | *P* Values Overall | $p \leq 0.01$ | $p \leq 0.05$ | $0.01 < p \leq 0.05$ | $0.03 < p \leq 0.05$ |
| --- | --- | --- | --- | --- | --- |
| **Statistical Power Increasing** | ↓ | ↑ | ↑ | ↓ | ↓ |
| ***P* Hacking Decreasing** | ↑ | ↓ | ↓ | ↓ | ↓ |
| ***P* Hacking Increasing** | ↓ | ↑ | ↑ | ↑ | ↑ |
| **Empirically Observed Trends** | ↓ | ↑ | ↑ | ↓ | ↓ |

When *P* hacking decreases and statistical power remains constant, regression coefficients indicate decreases in the total fractions less than or equal to 0.01 (per decade OR=0.8343) and decreases in the fractions less than or equal to 0.05 (per decade OR=0.2997), but increases in expected logit(*P* values). Decreases in P hacking also lead to decreases in *P* values at or just below the 0.05 threshold (e.g., 0.7922 per decade odds ratio for *P* values from 0.01 to 0.05).

Furthermore, decreases in *P* hacking as modeled by this strategy, lead to greater decreases in fraction of *P* values less than or equal to 0.05 (per decade OR=0.2997) than decreases seen in regions at or just below 0.05 (e.g., per annum OR=0.7922). Time trends of increasing P hacking induce the reverse pattern: increases in the fraction of P values below any threshold and increases in *P* values falling within a range just below 0.05. Table 6 qualitatively summarizes these results.

## Discussion

Using OpenAI’s API and ChatGPT’s 4o model, we derived *P* values from CIs in four major epidemiology journals from 2000-24. Logistic regression models showing declining odds of *P* values just below 0.05 alone could be misinterpreted as evidence that *P* hacking is decreasing, while the beta regression results showing overall declines in *P* values and logistic models showing the odds of a *P* value less than 0.05 or 0.01 are increasing complicate the picture.

*P* curve analyses clarify potential origins of observed trends. The observed distribution of *P* values over time is most consistent with the assumption that statistical power has increased since 2000. Increases in statistical power over time can produce increases in *P* values less than or equal to 0.01 and less than or equal to 0.05, concomitant with decreases in the intervals between 0.01 and 0.05 and between 0.03 and 0.05. These are precisely the trends observed in *P* values at major epidemiology journals.

Simulations from *P* curve models of decreasing *P* hacking over time with no changes in statistical power do not conform with the observed trends: *P* values overall in this scenario should be increasing rather than decreasing over time. Furthermore, simulations in which *P* hacking remains constant with no changes in statistical power failed to produce the empirical temporal trends in P values. These findings are most consistent with the conclusion that observed changes in the *P*-value distribution reflect increases in statistical power rather than decreased *P* hacking. Increases in statistical power are plausibly due to increases in sample sizes facilitated by the growing number of large, easily available cohort studies and electronic health record, genetic, and other sources of “big” data.

These results do not provide estimates of the prevalence of *P* hacking since results in abstracts are a selection of all analyses performed. Because of this, our analysis specifically looked at changes in the *P*-value distribution over time to infer changes in *P* hacking. Implicit in this is an assumption that the selection process from manuscript into abstract has remained more or less the same over time. However, the excess density observed just below the 0.05 threshold is likely indicative of *P* hacking and not only selection of results from the manuscript into the abstract. This is for two reasons: First, we observe excess density across the 0.01 to 0.05 range without excess density below 0.01 compared for all model fits. Second, we observe a blip in density just below 0.05 around 0.01 (Figure 1), patterns characteristic of *P* hacking.^29^ The precise mechanism by which this blip is produced in the epidemiologic literature is not clearly understood since our and others simulations of *P* hacking do not replicate this feature.^28^

Finally, our findings suggest the effect of declining *P* hacking on the distribution of *P* values would offset the effects of increasing statistical power. Thus, if *P* hacking is declining with time, the declines would have to be minimal, since such effects are more than compensated for by the effects of increasing statistical power. The simplest explanation is that we are seeing changes in the *P-*value distribution due solely or largely to changes in statistical power and not changes in *P* hacking.

*P* curves may have limited utility for detecting *P* hacking when it is present and the resulting *P* value distribution follows the characteristic J-shaped curve. A previous simulation study showed that the P value distribution under *P* hacking may be similar to the J-shaped distribution expected without P hacking (under an alternative hypothesis). This result contrasted with prior social psychology literature that assumed J-shaped *P* value distributions implied absence of P hacking.^28^ However, by fitting theoretical models, including mixture distributions, we showed that observed P value distributions have excess density just below the 0.05 threshold, deviating from expected J-shapes.

This analysis has the following strengths: First, our methods draw on and extend *P*-curve methods used in the social psychology literature to evaluate research credibility.^25–27^ Specifically, we model two-sided *P* value distributions and differentiate the cumulative distribution function to obtain a probability density function and directly fit this to data. Assuming all *P* values come from equally powered hypotheses can lead to incorrect conclusions,^30^ which motivated our fitting and reporting on multiple mixture models to determine which conclusions were robust to model assumptions. Second, we made use of sample splitting to avoid drawing conclusions about density in certain ranges using the same data that model fits were performed in.

This analysis has the following limitations: First, data extraction and *P*-value calculation procedures used produce approximate *P* values that are a subset of what is reported in journal articles. To maintain consistency in *P* values used in the analysis, all *P* values were calculated the same way whether or not an exact *P* value was reported. *P* values for a trend may be reported with contrasts of extreme *n*-tiles (e.g, the first and fourth quartile of an exposure). Thus, a *P* value appearing adjacent to an effect estimate and confidence interval may not correspond to this effect estimate and confidence interval. Additionally, we didn’t evaluate *P* values that were reported without confidence intervals. Based on prior literature^24^ and our qualitative assessment, *P* values reported without confidence intervals was rare even in the early 2000s.

Again, these *P* values were excluded to maintain consistency since they often qualitatively differ in purpose and importance in terms of overall results than quantities are reported with confidence intervals (e.g., *P* values for interactions or trends).^13^

In conclusion, we find that *P* values near the 0.05 threshold have decreased in epidemiology journals since 2000. Decreases in *P* hacking alone could not have produced the changes in *P* value distributions. Increases in sample sizes or other improvements in power since 2000 are consistent with observed trends in *P* value distributions from abstracts at epidemiology journals. The effects of the movement toward reducing reliance on *P* values and specific journal-level policies^1,13–16^ warrant empirical evaluation.

## Data Availability

All data produced are available on PubMed.

