## Supplemental Material for "Trends in the Distribution of *P Values* in Epidemiology Journals: Decreased *P* Hacking or Increased Power?"

Supplemental Tables
- Table S1: Summary of current policies related to the publication of P values at four major epidemiology journals.
- Table S2: Number of abstracts and confidence intervals and summary statistics by both journal and year.
- Table S3:  Regression coefficients from both beta and logistic regression fixed- and mixed-effects models stratified by journal.
- Table S3: Log Likelihoods and AICs of P-Curve Models.
- Table S4: Simulated fractions compared with fractions in the AJE for early and late times stratified at the median time.
- Median time is defined with respect to the confidence interval, not abstract and is 2011.4.
- Table S5: Simulated fractions compared with fractions in the AJE for early and late times stratified at 1/1/2012.
- Table S6: Simulated fractions compared with fractions in the AJE for early and late times stratified at 1/1/2016.
- Table S7. Comparison of automated to manual extraction of values in random subsamples by journal.

Supplemental Methods
- PubMed Search Strings
- Updated PubMed Search Strings
- Data Cleaning
- Justification for Including 0.05 in Intervals of Interest
- The Error Function

### Supplemental Tables

**Table S1:** Summary of current policies related to the publication of P values at four major epidemiology journals.

| Journal | Policies |
|---|---|
| Epidemiology | “Significance Testing: For estimates of causal effects, we strongly discourage the use of categorized P-values and language referring to statistical significance. We prefer interval estimation, which conveys the precision of the estimate with respect to sampling variability. We are more open to testing with respect to modeling decisions, such as for tests of interaction and for tests for trend, and with respect to studies using high-dimensional testing, such as genome-wide association or other genomic platforms.” |
| International Journal of Epidemiology | “In the IJE, we actively discourage the use of the term ‘statistically significant’ or just ‘significant’ and such statements in method sections as ‘findings at P<0.05 were considered significant’. Please provide effect estimates with confidence intervals and exact P values, and refrain from using the term ‘significant’ in either the results or discussion sections of papers.”<br><br>“Our justification of this position is given in: Sterne J, Davey-Smith G. Sifting the evidence — What’s wrong with significance tests? BMJ 2001; 322: 226-231. See also: Wasserstein RL, Lazar NA. The ASA’s statement on P-values: context, process, and purpose. The American Statistician 2016: DOI: 10.1080/00031305.2016.1154108.” |
| American Journal of Epidemiology | Implies that P values should only be reported in some situations, but does not specify what those situations are.<br>“*P* values: In cases in which *P* values should be reported, please note style for probability: *P* < 0.01, with an uppercase italic letter *P*. *P* values should not be bolded. Avoid reporting an excessive number of digits beyond the |

| | |
|---|---|
| | decimal for estimates, especially when the estimate has a wide confidence interval. If *P* values are given, they should be reported to, at most, 2 digits beyond any leading zeros. They may alternatively be reported as less than some specified value (e.g., $P < 0.05$ or $P < 0.001$). Indicate whether *P* values are 1 sided or 2 sided." |
| European Journal of Epidemiology | No guidelines given regarding *P* values. |
| CONSORT | Nothing explicit about P values, but there are instructions to report confidence intervals: "Outcomes and estimation: For each primary and secondary outcome, results for each group, and the estimated effect size and its precision (such as 95% confidence interval)." |
| STROBE<br>https://www.strobe-statement.org/ | Nothing explicit about P values, but there are instructions to report confidence intervals: "Give unadjusted estimates and, if applicable, confounder-adjusted estimates and their precision (eg, 95% confidence interval)." |
| PRISMA<br>http://www.prisma-statement.org/ | Nothing explicit about P values, but there are instructions to report confidence intervals: "For all outcomes, present, for each study: (a) summary statistics for each group (where appropriate) and (b) an effect estimate and its precision (e.g. confidence/credible interval), ideally using structured tables or plots."<br><br>"Present results of all statistical syntheses conducted. If meta-analysis was done, present for each the summary estimate and its precision (e.g. confidence/credible interval) and measures of statistical heterogeneity. If comparing groups, describe the direction of the effect." |

**Table S2:** Number of abstracts and confidence intervals and summary statistics by both journal and year.

Epi=*Epidemiology*, AJE=*American Journal of Epidemiology*, EJE=*European Journal of Epidemiology*, IJE=*International Journal of Epidemiology.* IQR: interquartile range; N: number; CI: confidence interval.

| Journal | Year | N of Abstracts | N of CIs | Mean of P | Median of log P (IQR) |
|---|---|---|---|---|---|
| Epi | 2000 | 60 | 178 | 0.113 | -3.82, (-7.2, -2.4) |
| Epi | 2001 | 70 | 238 | 0.107 | -3.94, (-9.46, -2.27) |
| Epi | 2002 | 50 | 152 | 0.132 | -4.08, (-6.88, -2.27) |
| Epi | 2003 | 58 | 149 | 0.15 | -4.27, (-7.19, -2.37) |
| Epi | 2004 | 57 | 172 | 0.127 | -3.98, (-6.71, -2.27) |
| Epi | 2005 | 63 | 187 | 0.104 | -4.01, (-8.08, -2.4) |
| Epi | 2006 | 52 | 157 | 0.116 | -3.98, (-7.37, -2.85) |
| Epi | 2007 | 54 | 182 | 0.0818 | -4.97, (-8.93, -3.2) |
| Epi | 2008 | 52 | 162 | 0.13 | -4.08, (-8.99, -2.21) |
| Epi | 2009 | 64 | 204 | 0.124 | -4.28, (-9.94, -2.56) |
| Epi | 2010 | 55 | 162 | 0.0908 | -4.89, (-23.4, -2.9) |
| Epi | 2011 | 56 | 180 | 0.0912 | -4.74, (-11.6, -2.9) |
| Epi | 2012 | 56 | 192 | 0.0765 | -5.19, (-13.6, -2.99) |
| Epi | 2013 | 51 | 168 | 0.117 | -4.5, (-9.69, -2.74) |
| Epi | 2014 | 58 | 174 | 0.076 | -5.25, (-12.6, -3.3) |
| Epi | 2015 | 68 | 220 | 0.11 | -5.43, (-13.6, -2.93) |
| Epi | 2016 | 53 | 152 | 0.119 | -4.01, (-9.77, -2.3) |
| Epi | 2017 | 69 | 208 | 0.0847 | -7.52, (-33.6, -3.47) |
| Epi | 2018 | 67 | 237 | 0.0666 | -6.78, (-24.6, -3.59) |

| | | | | | |
|---|---|---|---|---|---|
| Epi | 2019 | 73 | 246 | 0.0864 | -5.57, (-29.5, -3.13) |
| Epi | 2020 | 55 | 189 | 0.0703 | -11, (-114, -4.27) |
| Epi | 2021 | 54 | 170 | 0.0955 | -6.08, (-32.7, -3.05) |
| Epi | 2022 | 50 | 141 | 0.0726 | -10.4, (-97, -3.67) |
| Epi | 2023 | 39 | 115 | 0.145 | -5.25, (-20.5, -1.81) |
| Epi | 2024 | 32 | 106 | 0.0835 | -9.09, (-23.9, -3.2) |
| AJE | 2000 | 149 | 531 | 0.107 | -4.54, (-9.19, -2.84) |
| AJE | 2001 | 125 | 389 | 0.0947 | -4.3, (-7.39, -2.7) |
| AJE | 2002 | 121 | 385 | 0.125 | -4.38, (-9.36, -2.65) |
| AJE | 2003 | 146 | 500 | 0.117 | -4.35, (-7.92, -2.3) |
| AJE | 2004 | 165 | 488 | 0.0986 | -4.7, (-8.36, -2.88) |
| AJE | 2005 | 141 | 436 | 0.101 | -4.74, (-9.99, -3.02) |
| AJE | 2006 | 179 | 573 | 0.0912 | -4.82, (-9.48, -3.05) |
| AJE | 2007 | 190 | 540 | 0.0878 | -4.86, (-10.8, -3.15) |
| AJE | 2008 | 178 | 540 | 0.0882 | -4.69, (-10.9, -3.11) |
| AJE | 2009 | 173 | 535 | 0.0923 | -5.11, (-10.1, -3.2) |
| AJE | 2010 | 151 | 414 | 0.101 | -5, (-10.2, -3.16) |
| AJE | 2011 | 140 | 399 | 0.0819 | -5.65, (-12.6, -3.28) |
| AJE | 2012 | 169 | 498 | 0.0774 | -5.61, (-15, -3.34) |
| AJE | 2013 | 194 | 534 | 0.0759 | -5.54, (-16.5, -3.18) |
| AJE | 2014 | 118 | 332 | 0.0645 | -6.37, (-15.5, -3.45) |
| AJE | 2015 | 111 | 328 | 0.0965 | -5.15, (-10.1, -3.17) |
| AJE | 2016 | 114 | 338 | 0.0696 | -6.15, (-17.4, -3.5) |
| AJE | 2017 | 161 | 514 | 0.0715 | -6.55, (-23, -3.51) |

| | | | | | |
|---|---|---|---|---|---|
| AJE | 2018 | 128 | 388 | 0.0574 | -6.38, (-21.6, -3.9) |
| AJE | 2019 | 78 | 246 | 0.0795 | -5.17, (-12.4, -3.38) |
| AJE | 2020 | 110 | 349 | 0.121 | -5.54, (-18.9, -3.1) |
| AJE | 2021 | 111 | 317 | 0.0734 | -6.75, (-35.5, -3.56) |
| AJE | 2022 | 63 | 189 | 0.0565 | -7.39, (-38, -3.72) |
| AJE | 2023 | 78 | 221 | 0.0658 | -6.66, (-36, -3.4) |
| AJE | 2024 | 134 | 435 | 0.0809 | -5.37, (-16.9, -3.22) |
| EJE | 2000 | 18 | 49 | 0.0548 | -10.9, (-88.8, -4.54) |
| EJE | 2001 | 42 | 123 | 0.0134 | -13.7, (-77.3, -4.97) |
| EJE | 2003 | 28 | 84 | 0.0699 | -4.35, (-7.04, -3.1) |
| EJE | 2004 | 37 | 112 | 0.0409 | -5.61, (-19.5, -3.41) |
| EJE | 2005 | 47 | 172 | 0.0812 | -5.32, (-17.4, -3.21) |
| EJE | 2006 | 39 | 129 | 0.0772 | -4.22, (-6.96, -3.21) |
| EJE | 2007 | 41 | 143 | 0.0679 | -5.41, (-16.2, -3.18) |
| EJE | 2008 | 38 | 123 | 0.0738 | -7.32, (-13.7, -3.5) |
| EJE | 2009 | 45 | 137 | 0.0898 | -6.94, (-24.9, -3.57) |
| EJE | 2010 | 53 | 170 | 0.07 | -5.93, (-17.5, -3.54) |
| EJE | 2011 | 43 | 142 | 0.0939 | -5.33, (-15.2, -3.14) |
| EJE | 2012 | 58 | 220 | 0.0994 | -5, (-13.2, -2.89) |
| EJE | 2013 | 51 | 180 | 0.0859 | -5.43, (-14.2, -3.08) |
| EJE | 2014 | 43 | 170 | 0.075 | -4.71, (-9, -3.03) |
| EJE | 2015 | 49 | 191 | 0.086 | -5, (-11.8, -3.21) |
| EJE | 2016 | 55 | 206 | 0.074 | -6.77, (-15.5, -3.15) |
| EJE | 2017 | 55 | 182 | 0.073 | -5.6, (-14.6, -3.39) |

| | | | | | |
|---|---|---|---|---|---|
| EJE | 2018 | 61 | 271 | 0.0423 | -8.29, (-33, -4.16) |
| EJE | 2019 | 49 | 167 | 0.118 | -6.03, (-15.4, -3.15) |
| EJE | 2020 | 48 | 170 | 0.074 | -7.1, (-29, -3.46) |
| EJE | 2021 | 61 | 236 | 0.0581 | -7.89, (-30.4, -4.02) |
| EJE | 2022 | 53 | 206 | 0.0908 | -6.45, (-23.8, -3.58) |
| EJE | 2023 | 45 | 175 | 0.0781 | -5.61, (-11.6, -3.44) |
| EJE | 2024 | 24 | 99 | 0.0713 | -9.45, (-44.8, -3.6) |
| IJE | 2000 | 47 | 161 | 0.0646 | -5.19, (-13, -3.06) |
| IJE | 2001 | 58 | 219 | 0.0891 | -5.37, (-16.9, -3.16) |
| IJE | 2002 | 48 | 141 | 0.0803 | -4.72, (-10.4, -3.18) |
| IJE | 2003 | 48 | 150 | 0.0662 | -5.29, (-11.6, -3.29) |
| IJE | 2004 | 52 | 201 | 0.0733 | -7.39, (-17.9, -3.84) |
| IJE | 2005 | 56 | 216 | 0.0418 | -7.61, (-23.6, -4.06) |
| IJE | 2006 | 62 | 223 | 0.0997 | -5.4, (-19.4, -3.07) |
| IJE | 2007 | 48 | 163 | 0.0504 | -5.58, (-11.3, -3.69) |
| IJE | 2008 | 59 | 206 | 0.067 | -5.23, (-12.7, -3.4) |
| IJE | 2009 | 75 | 240 | 0.0637 | -6.11, (-16.4, -3.47) |
| IJE | 2010 | 65 | 231 | 0.0929 | -5.27, (-12.9, -2.92) |
| IJE | 2011 | 64 | 245 | 0.0816 | -5.31, (-12.3, -3.29) |
| IJE | 2012 | 53 | 186 | 0.0629 | -6.33, (-14.1, -3.73) |
| IJE | 2013 | 53 | 185 | 0.0863 | -7.88, (-28.8, -3.4) |
| IJE | 2014 | 63 | 290 | 0.0847 | -6.92, (-26.9, -3.41) |
| IJE | 2015 | 43 | 164 | 0.0645 | -6.23, (-23.5, -3.56) |
| IJE | 2016 | 103 | 398 | 0.0912 | -5.22, (-14.4, -3.03) |

| | | | | | |
|---|---|---|---|---|---|
| IJE | 2017 | 104 | 351 | 0.105 | -6.34, (-19.9, -3.31) |
| IJE | 2018 | 95 | 306 | 0.0664 | -8.69, (-25.5, -3.59) |
| IJE | 2019 | 102 | 385 | 0.079 | -6.67, (-40.7, -3.29) |
| IJE | 2020 | 93 | 335 | 0.0745 | -8.3, (-38.8, -3.97) |
| IJE | 2021 | 95 | 346 | 0.0796 | -7.48, (-41.8, -3.56) |
| IJE | 2022 | 100 | 432 | 0.0598 | -8.25, (-31, -4.02) |
| IJE | 2023 | 112 | 448 | 0.0488 | -8.38, (-26.4, -4.27) |
| IJE | 2024 | 22 | 83 | 0.0198 | -22.4, (-52.3, -7.32) |
| IJE | 2024 | 22 | 83 | 0.0198 | -22.4, (-52.3, -7.32) |

**Table S3:** Regression coefficients from both beta and logistic regression fixed- and mixed-effects models stratified by journal.

| | Fixed-Effects Models | Mixed-Effects Models |
|---|---|---|
| **Beta Regression** $\exp(E[logit(p)])$ | | |
| Epidemiology | 0.192, 95% CI: (0.185, 0.199) | 0.1687, 95% CI: (0.1594, 0.1786) |
| AJE | 0.182, 95% CI: (0.177, 0.188) | 0.1534, 95% CI: (0.1407, 0.1671) |
| EJE | 0.161, 95% CI: (0.155, 0.168) | 0.1303, 95% CI: (0.1179, 0.1441) |
| IJE | 0.156, 95% CI: (0.151, 0.162) | 0.1256, 95% CI: (0.1145, 0.1377) |
| **Beta Regression Per-decade ratio of** $E[p/(1 - E[p])$ | | |
| Epidemiology | 0.746, 95% CI: (0.7115, 0.7823) | 0.7552, 95% CI: (0.7005, 0.8142) |
| AJE | 0.8142, 95% CI: (0.789, 0.8402) | 0.8148, 95% CI: (0.7242, 0.9167) |
| EJE | 0.978, 95% CI: (0.927, 1.032) | 1.018, 95% CI: (0.8861, 1.17) |
| IJE | 0.8823, 95% CI: (0.8484, 0.9175) | 0.8822, 95% CI: (0.7783, 1) |
| **Logistic Regression: Per-decade odds ratio** $p \in (0, 0.01]$ | | |
| Epidemiology | 1.549, 95% CI: (1.425, 1.685) | 1.845, 95% CI: (1.578, 2.157) |
| AJE | 1.341, 95% CI: (1.268, 1.418) | 1.484, 95% CI: (1.163, 1.894) |
| EJE | 1.09, 95% CI: (0.9898, 1.201) | 1.076, 95% CI: (0.8059, 1.436) |
| IJE | 1.237, 95% CI: (1.152, 1.329) | 1.339, 95% CI: (1.032, 1.737) |
| **Logistic Regression: Per-decade odds ratio** $p \in (0, 0.05]$ | | |
| Epidemiology | 1.378, 95% CI: (1.256, 1.512) | 1.546, 95% CI: (1.303, 1.834) |
| AJE | 1.32, 95% CI: (1.235, 1.411) | 1.437, 95% CI: (1.098, 1.88) |
| EJE | 0.9497, 95% CI: (0.8422, 1.071) | 0.872, 95% CI: (0.6294, 1.208) |

| | | |
|---|---|---|
| IJE | 1.164, 95% CI: (1.067, 1.27) | 1.258, 95% CI: (0.9415, 1.682) |
| **Logistic Regression: Per-decade odds ratio** $p \in (0.01, 0.05]$ | | |
| Epidemiology | 0.7617, 95% CI: (0.6879, 0.8435) | 0.7281, 95% CI: (0.6327, 0.8378) |
| AJE | 0.8695, 95% CI: (0.8134, 0.9294) | 0.8386, 95% CI: (0.6736, 1.044) |
| EJE | 0.8365, 95% CI: (0.7437, 0.9408) | 0.8157, 95% CI: (0.6299, 1.056) |
| IJE | 0.8394, 95% CI: (0.7679, 0.9175) | 0.8231, 95% CI: (0.6511, 1.04) |
| **Logistic Regression: Per-decade odds ratio** $p \in (0.03, 0.05]$ | | |
| Epidemiology | 0.8537, 95% CI: (0.7303, 0.998) | 0.8346, 95% CI: (0.6836, 1.019) |
| AJE | 0.8636, 95% CI: (0.7789, 0.9575) | 0.8409, 95% CI: (0.6155, 1.149) |
| EJE | 0.8328, 95% CI: (0.6932, 1.001) | 0.8182, 95% CI: (0.565, 1.185) |
| IJE | 0.8715, 95% CI: (0.7602, 0.999) | 0.8658, 95% CI: (0.6204, 1.208) |

**Table S3:** Log Likelihoods and AICs of P-Curve Models.

Models are as follows: 1. One standardized effect size ($\theta$); 2. One standardized effect size ($\theta$), linear trend; 3. Two standardized effect sizes ($\theta_1$=0 and $\theta_2$); 4. Two standardized effect sizes ($\theta_1$=0 and $\theta_2$), linear trend; 5. Exponential mixture; 6. Exponential mixture, linear trend; 7. 8. Gamma mixture; and 8. Gamma mixture, linear trend.

| **Model** | **Log Likelihood** | **Number of Parameters** | **AIC** |
|---|---|---|---|
| 1 | 63599.5 | 1 | -127197 |
| 2 | 63997.4 | 2 | -127990.7 |
| 3 | 68038.8 | 2 | -136073.6 |
| 4 | 68421.3 | 3 | -136836.7 |
| 5 | 71564.8 | 1 | -143127.6 |
| 6 | 71606 | 2 | -143207.9 |
| 7 | 73555.4 | 2 | -147106.8 |
| 8 | 73654.2 | 3 | -147302.4 |

**Table S4:** Simulated fractions compared with fractions in the AJE for early and late times stratified at the median time.

Median time is defined with respect to the confidence interval, not abstract and is 2011.4.

| | **Mean Fraction** | **Minimum Fraction** | **Maximum Fraction** | **Fraction in Empirical Data** |
|---|---|---|---|---|
| **Model 2** | | | | |
| Early: Between 0.01 and 0.05 | 0.167 | 0.155 | 0.177 | 0.221 |
| Late: Between 0.01 and 0.05 | 0.105 | 0.097 | 0.115 | 0.18 |
| Early: Between 0.03 and 0.05 | 0.047 | 0.042 | 0.056 | 0.078 |
| Late: Between 0.03 and 0.05 | 0.027 | 0.022 | 0.031 | 0.064 |
| Early: Over 0.05 | 0.12 | 0.109 | 0.132 | 0.243 |
| Late: Over 0.05 | 0.053 | 0.048 | 0.06 | 0.189 |
| Early: Less than 0.01 | 0.713 | 0.701 | 0.725 | 0.536 |
| Late: Less than 0.01 | 0.842 | 0.833 | 0.852 | 0.631 |
| Early: All less than 0.05 | 0.88 | 0.868 | 0.891 | 0.757 |
| Late: All less than 0.05 | 0.947 | 0.94 | 0.952 | 0.811 |
| **Model 4** | | | | |
| Early: Between 0.01 and 0.05 | 0.164 | 0.147 | 0.174 | 0.221 |
| Late: Between 0.01 and 0.05 | 0.121 | 0.112 | 0.132 | 0.18 |
| Early: Between 0.03 and 0.05 | 0.052 | 0.045 | 0.058 | 0.078 |

| | | | | |
|---|---|---|---|---|
| Late: Between 0.03 and 0.05 | 0.035 | 0.03 | 0.042 | 0.064 |
| Early: Over 0.05 | 0.386 | 0.375 | 0.4 | 0.243 |
| Late: Over 0.05 | 0.311 | 0.296 | 0.327 | 0.189 |
| Early: Less than 0.01 | 0.451 | 0.435 | 0.471 | 0.536 |
| Late: Less than 0.01 | 0.569 | 0.555 | 0.581 | 0.631 |
| Early: All less than 0.05 | 0.614 | 0.6 | 0.625 | 0.757 |
| Late: All less than 0.05 | 0.689 | 0.673 | 0.704 | 0.811 |
| **Model 6** | | | | |
| Early: Between 0.01 and 0.05 | 0.103 | 0.092 | 0.113 | 0.221 |
| Late: Between 0.01 and 0.05 | 0.098 | 0.089 | 0.106 | 0.18 |
| Early: Between 0.03 and 0.05 | 0.037 | 0.032 | 0.044 | 0.078 |
| Late: Between 0.03 and 0.05 | 0.035 | 0.03 | 0.039 | 0.064 |
| Early: Over 0.05 | 0.46 | 0.444 | 0.475 | 0.243 |
| Late: Over 0.05 | 0.418 | 0.407 | 0.429 | 0.189 |
| Early: Less than 0.01 | 0.437 | 0.417 | 0.452 | 0.536 |
| Late: Less than 0.01 | 0.485 | 0.473 | 0.5 | 0.631 |
| Early: All less than 0.05 | 0.54 | 0.525 | 0.556 | 0.757 |
| Late: All less than 0.05 | 0.582 | 0.571 | 0.593 | 0.811 |
| **Model 8** | | | | |
| Early: Between 0.01 and 0.05 | 0.133 | 0.124 | 0.146 | 0.221 |
| Late: Between 0.01 and 0.05 | 0.119 | 0.11 | 0.131 | 0.18 |

| | | | | |
|---|---|---|---|---|
| Early: Between 0.03 and 0.05 | 0.045 | 0.038 | 0.051 | 0.078 |
| Late: Between 0.03 and 0.05 | 0.038 | 0.034 | 0.045 | 0.064 |
| Early: Over 0.05 | 0.295 | 0.282 | 0.304 | 0.243 |
| Late: Over 0.05 | 0.224 | 0.21 | 0.235 | 0.189 |
| Early: Less than 0.01 | 0.572 | 0.557 | 0.59 | 0.536 |
| Late: Less than 0.01 | 0.658 | 0.644 | 0.671 | 0.631 |
| Early: All less than 0.05 | 0.705 | 0.696 | 0.718 | 0.757 |
| Late: All less than 0.05 | 0.776 | 0.765 | 0.79 | 0.811 |

**Table S5:** Simulated fractions compared with fractions in the AJE for early and late times stratified at 1/1/2012.

Models are as follows: 2. One standardized effect size ($\theta$), linear trend; 4. Two standardized effect sizes ($\theta_1$=0 and $\theta_2$), linear trend; 6. Exponential mixture, linear trend; and 8. Gamma mixture, linear trend. Model parameters from fits to the AJE testing set are used, while simulation abstract dates and the fractions in AJE are from the testing set.

| | **Mean Fraction** | **Minimum Fraction** | **Maximum Fraction** | **Fraction in Empierical Data** |
|---|---|---|---|---|
| **Model 2** | | | | |
| Early: Between 0.01 and 0.05 | 0.168 | 0.156 | 0.178 | 0.223 |
| Late: Between 0.01 and 0.05 | 0.106 | 0.099 | 0.116 | 0.18 |
| Early: Between 0.03 and 0.05 | 0.048 | 0.042 | 0.057 | 0.079 |
| Late: Between 0.03 and 0.05 | 0.027 | 0.023 | 0.031 | 0.063 |
| Early: Over 0.05 | 0.122 | 0.11 | 0.133 | 0.244 |
| Late: Over 0.05 | 0.054 | 0.049 | 0.06 | 0.19 |
| Early: Less than 0.01 | 0.71 | 0.698 | 0.724 | 0.533 |
| Late: Less than 0.01 | 0.84 | 0.83 | 0.849 | 0.63 |
| Early: All less than 0.05 | 0.878 | 0.867 | 0.89 | 0.756 |
| Late: All less than 0.05 | 0.946 | 0.94 | 0.951 | 0.81 |
| **Model 4** | | | | |
| Early: Between 0.01 and 0.05 | 0.164 | 0.147 | 0.175 | 0.223 |
| Late: Between 0.01 and 0.05 | 0.122 | 0.113 | 0.133 | 0.18 |

| | | | | |
|---|---|---|---|---|
| Early: Between 0.03 and 0.05 | 0.052 | 0.045 | 0.059 | 0.079 |
| Late: Between 0.03 and 0.05 | 0.036 | 0.031 | 0.043 | 0.063 |
| Early: Over 0.05 | 0.388 | 0.375 | 0.402 | 0.244 |
| Late: Over 0.05 | 0.312 | 0.296 | 0.326 | 0.19 |
| Early: Less than 0.01 | 0.448 | 0.433 | 0.469 | 0.533 |
| Late: Less than 0.01 | 0.567 | 0.554 | 0.579 | 0.63 |
| Early: All less than 0.05 | 0.612 | 0.598 | 0.625 | 0.756 |
| Late: All less than 0.05 | 0.688 | 0.674 | 0.704 | 0.81 |
| **Model 6** | | | | |
| Early: Between 0.01 and 0.05 | 0.103 | 0.094 | 0.115 | 0.223 |
| Late: Between 0.01 and 0.05 | 0.098 | 0.09 | 0.106 | 0.18 |
| Early: Between 0.03 and 0.05 | 0.037 | 0.031 | 0.044 | 0.079 |
| Late: Between 0.03 and 0.05 | 0.035 | 0.029 | 0.04 | 0.063 |
| Early: Over 0.05 | 0.461 | 0.446 | 0.475 | 0.244 |
| Late: Over 0.05 | 0.418 | 0.405 | 0.43 | 0.19 |
| Early: Less than 0.01 | 0.436 | 0.418 | 0.452 | 0.533 |
| Late: Less than 0.01 | 0.484 | 0.472 | 0.5 | 0.63 |
| Early: All less than 0.05 | 0.539 | 0.525 | 0.554 | 0.756 |
| Late: All less than 0.05 | 0.582 | 0.57 | 0.595 | 0.81 |
| **Model 8** | | | | |
| Early: Between 0.01 and 0.05 | 0.134 | 0.124 | 0.147 | 0.223 |

| | | | | |
|---|---|---|---|---|
| Late: Between 0.01 and 0.05 | 0.119 | 0.111 | 0.131 | 0.18 |
| Early: Between 0.03 and 0.05 | 0.045 | 0.038 | 0.051 | 0.079 |
| Late: Between 0.03 and 0.05 | 0.038 | 0.034 | 0.044 | 0.063 |
| Early: Over 0.05 | 0.297 | 0.282 | 0.308 | 0.244 |
| Late: Over 0.05 | 0.225 | 0.211 | 0.236 | 0.19 |
| Early: Less than 0.01 | 0.57 | 0.552 | 0.59 | 0.533 |
| Late: Less than 0.01 | 0.656 | 0.644 | 0.669 | 0.63 |
| Early: All less than 0.05 | 0.703 | 0.692 | 0.718 | 0.756 |
| Late: All less than 0.05 | 0.775 | 0.764 | 0.789 | 0.81 |

**Table S6:** Simulated fractions compared with fractions in the AJE for early and late times stratified at 1/1/2016.

Models are as follows: 2. One standardized effect size ($z$), linear trend; 4. Two standardized effect sizes ($\theta_1$=0 and $\theta_2$), linear trend; 6. Exponential mixture, linear trend; and 8. Gamma mixture, linear trend. Model parameters from fits to the AJE testing set are used, while simulation abstract dates and the fractions in AJE are from the testing set.

| | **Mean Fraction** | **Minimum Fraction** | **Maximum Fraction** | **Fraction in Empirical Data** |
|---|---|---|---|---|
| **Model 2** | | | | |
| Early: Between 0.01 and 0.05 | 0.158 | 0.15 | 0.167 | 0.216 |
| Late: Between 0.01 and 0.05 | 0.097 | 0.085 | 0.106 | 0.173 |
| Early: Between 0.03 and 0.05 | 0.044 | 0.039 | 0.051 | 0.076 |
| Late: Between 0.03 and 0.05 | 0.024 | 0.02 | 0.03 | 0.063 |
| Early: Over 0.05 | 0.109 | 0.101 | 0.12 | 0.238 |
| Late: Over 0.05 | 0.047 | 0.038 | 0.056 | 0.179 |
| Early: Less than 0.01 | 0.733 | 0.72 | 0.742 | 0.546 |
| Late: Less than 0.01 | 0.856 | 0.845 | 0.867 | 0.648 |
| Early: All less than 0.05 | 0.891 | 0.88 | 0.899 | 0.762 |
| Late: All less than 0.05 | 0.953 | 0.944 | 0.962 | 0.821 |
| **Model 4** | | | | |
| Early: Between 0.01 and 0.05 | 0.158 | 0.145 | 0.168 | 0.216 |
| Late: Between 0.01 and 0.05 | 0.114 | 0.104 | 0.124 | 0.173 |

| | | | | |
|---|---|---|---|---|
| Early: Between 0.03 and 0.05 | 0.049 | 0.045 | 0.055 | 0.076 |
| Late: Between 0.03 and 0.05 | 0.033 | 0.027 | 0.04 | 0.063 |
| Early: Over 0.05 | 0.374 | 0.363 | 0.385 | 0.238 |
| Late: Over 0.05 | 0.303 | 0.285 | 0.323 | 0.179 |
| Early: Less than 0.01 | 0.468 | 0.454 | 0.484 | 0.546 |
| Late: Less than 0.01 | 0.583 | 0.565 | 0.6 | 0.648 |
| Early: All less than 0.05 | 0.626 | 0.615 | 0.637 | 0.762 |
| Late: All less than 0.05 | 0.697 | 0.677 | 0.715 | 0.821 |
| **Model 6** | | | | |
| Early: Between 0.01 and 0.05 | 0.102 | 0.093 | 0.109 | 0.216 |
| Late: Between 0.01 and 0.05 | 0.097 | 0.087 | 0.105 | 0.173 |
| Early: Between 0.03 and 0.05 | 0.037 | 0.033 | 0.043 | 0.076 |
| Late: Between 0.03 and 0.05 | 0.035 | 0.029 | 0.041 | 0.063 |
| Early: Over 0.05 | 0.454 | 0.439 | 0.467 | 0.238 |
| Late: Over 0.05 | 0.413 | 0.392 | 0.427 | 0.179 |
| Early: Less than 0.01 | 0.444 | 0.429 | 0.456 | 0.546 |
| Late: Less than 0.01 | 0.491 | 0.475 | 0.508 | 0.648 |
| Early: All less than 0.05 | 0.546 | 0.533 | 0.561 | 0.762 |
| Late: All less than 0.05 | 0.587 | 0.573 | 0.608 | 0.821 |
| **Model 8** | | | | |
| Early: Between 0.01 and 0.05 | 0.131 | 0.121 | 0.143 | 0.216 |

| | | | | |
|---|---|---|---|---|
| Late: Between 0.01 and 0.05 | 0.117 | 0.108 | 0.128 | 0.173 |
| Early: Between 0.03 and 0.05 | 0.044 | 0.037 | 0.049 | 0.076 |
| Late: Between 0.03 and 0.05 | 0.038 | 0.031 | 0.045 | 0.063 |
| Early: Over 0.05 | 0.285 | 0.274 | 0.297 | 0.238 |
| Late: Over 0.05 | 0.215 | 0.2 | 0.23 | 0.179 |
| Early: Less than 0.01 | 0.584 | 0.571 | 0.596 | 0.546 |
| Late: Less than 0.01 | 0.669 | 0.65 | 0.688 | 0.648 |
| Early: All less than 0.05 | 0.715 | 0.703 | 0.726 | 0.762 |
| Late: All less than 0.05 | 0.785 | 0.77 | 0.8 | 0.821 |

**Table S7.** Comparison of automated to manual extraction of values in random subsamples by journal.

| Journal | Epidemiology | AJE | EJE | IJE |
|---|---|---|---|---|
| **abstracts sampled from LLM output** | 25 | 25 | 25 | 25 |
| **abstracts containing CIs** | 13 | 16 | 12 | 15 |
| **total CIs extracted** | 34 | 45 | 42 | 45 |
| **total CIs incorrect** | 0 | 0 | 3 | 2 |
| **total missing from LLM output** | 0 | 0 | 0 | 3 |
| **% entries containing error** | 0.00 | 0.00 | 7.14 | 4.4 |
| **95% CI (% error)** | 0 - 10.2 | 0 - 7.8 | 1.5 - 19.4 | 0.5 - 15.1 |

### Supplemental Methods

**PubMed Search Strings**

"Epidemiology"[Journal] AND 2000/01/01:2024/11/05[Date - Publication]

"American Journal of Epidemiology"[Journal] AND 2000/01/01:2024/12/31[Date - Publication]

"European Journal of Epidemiology"[Journal] AND 2000/01/01:2024/12/31[Date - Publication]

"International Journal of Epidemiology"[Journal] AND 2000/01/01:2024/12/31[Date - Publication]

**Data Cleaning**

Abstracts with missing day of the month of publication were assigned the 15th of the month; abstracts with missing day and month were assigned June 15th.

*P* values less than $10^{-10}$ were assigned a value of $10^{-10}$ and greater than $1 - 10^{-10}$ were assigned a value of $1 - 10^{-10}$.

**Justification for Including 0.05 in Intervals of Interest**

Contrary to convention, we considered *P* values equal to 0.05 in these ranges since confidence intervals that do not contain the null may produce a derived *P* value of 0.05 due to rounding in the original abstract. For example, if a rounded effect estimate of 0.3 and rounded 95% CI (0.0, 0.6) is given, the calculated *P* value would be exactly 0.05. However, this is more likely to correspond to a *P* value just under 0.05 due to both the steep shape of *P* curves and the presumed presence of *P* hacking.

#### The Error Function

$$erf(x) = \frac{2}{\sqrt{\pi}} \int_{\infty}^{x} e^{-t^2} dt$$
